# Gaps in Artificial Intelligence Research for Rural Health in the United States: A Scoping Review

**DOI:** 10.1101/2025.06.26.25330361

**Authors:** Katherine E. Brown, Sharon E. Davis

**Affiliations:** Department of Biomedical Informatics, Vanderbilt University Medical Center, Nashville, TN

**Keywords:** artificial intelligence, machine learning, rural US healthcare

## Abstract

**Background:** Artificial intelligence (AI) has impacted healthcare at urban and academic medical centers globally. The current focus on AI deployments in urban areas and the history of US urban-rural digital divides raises concerns that the promise of AI may not be realized in rural communities. This may exacerbate well-documented health disparities. Without the benefits of AI-driven improvements in patient outcomes and increased efficiency, rural healthcare facilities may fall farther behind their urban counterparts and rural hospital closure rates may continue to rise.

**Methods:** We conducted a scoping review following the PRISMA guidelines. We included peer-reviewed, original research studies indexed in PubMed, Embase, and WebOfScience after January 1, 2010 and through April 29, 2025. Studies were required to discuss the development, implementation, or evaluation of AI tools in rural US healthcare, including frameworks that help facilitate AI development (e.g., data warehouses).

**Findings:** Our search strategy found 26 studies meeting inclusion criteria after full text screening with 14 papers discussing predictive AI models and 12 papers discussing data or research infrastructure. AI models most commonly targeted resource allocation and distribution. Few studies explored model deployment and impact. Half noted the lack of data and analytic resources as a limitation to both development and validation. None of the studies discussed examples of generative AI being trained, evaluated, or deployed in a rural setting.

**Interpretation:** Practical limitations may be influencing and limiting the types of AI models evaluated in the rural US. We noted validation of tools in the rural US was underwhelming, and ultimately, neglected. With few studies moving beyond AI model design and development stages, there is a clear gap in our understanding of how to reliably validate, deploy, and sustain AI models in rural settings to advance health in all communities.

**Funding:** National Library of Medicine

**Research in context:** Evidence before this study: Clinical artificial intelligence (AI)—both for prediction modeling and generative tools— tools promise to reduce care delays, improve diagnosis and treatment decision-making, reduce care costs, and improve efficiency to reduce provider workload and enhance practice management. Unfortunately, efforts to deploy artificial intelligence (AI)—both for prediction modeling and generative tools—in healthcare are advancing, primarily at large academic medical centers and in urban areas. An emerging new digital divide in the use of clinical AI could exacerbate the well-documented health disparities between urban and rural communities in the United States. A better understanding of if and how AI is being developed, deployed, and evaluated across rural US communities is necessary to identify resources gaps and challenges to broad AI use in all communities.

Added value of this study: This study analyzes the current state of artificial intelligence research in the rural United States. For predictive AI models, applications most commonly targeted resource allocation and distribution. We noted several attempts to predict resource utilization of patients who were either tested or tested positive to COVID-19. However, we noted few AI solutions for acute medical events faced by rural patients, such as trauma and stroke, despite worse outcomes for rural patients suffering from these acute events. The limited availability of time-critical specialties such as trauma/emergency medicine, neurology, and cardiology in rural areas often necessitates patients with such conditions be transferred to larger, more resourced hospitals. Practical limitations may be influencing and limiting the types of AI models evaluated in rural US medical facilities. The most frequent model employed were tree-based ensembles, such as random forests and gradient-boosting trees. Our review also highlighted few studies of AI models moving beyond the design and develop stages, leaving a clear gap in our understanding of how to deploy and sustain predictive AI models in rural settings. Several challenges noted in the reviewed studies may provide insight into this lack of translation from research to implementation. We note that validation of A tools in the rural US was underwhelming, and ultimately, neglected. The most common form of model validation employed was a single random holdout test set. Half of the included papers mentioned a lack of reliable data sources or limited data volume as a potential challenge in developing and adopting AI/ML tools. The use of patient-level EHR data was often limited to what was available to the authors or at a specific medical center.

Implications of all the available evidence: Our review indicates a gap and highlights opportunity for innovation in leveraging AI tools to predict and support patients in rural communities. Further research is needed to enhance the translation of state-of-the-art modeling techniques into effective AI tools for use in the rural US, including exploring partnerships between academic medical centers and rural communities and solutions to logistic challenges of such partnerships, including data and resource sharing.

## Introduction

Artificial intelligence (AI) – herein defined as discriminative models such as machine learning (ML) *and* generative models such as large language models (LLMs) – has impacted healthcare at urban and academic medical centers globally. In São Paulo, Brazil, a hospital implemented an AI tool to streamline patient registrations, reducing wait times and increasing provider time with patients during the COVID-19 pandemic (1). In China, AI tools to predict public health trends to more efficiently allocate resources and target interventions in urban areas of central and eastern regions of the country (2). Several studies in European medical centers have documented radiology use cases in which AI reduced read times and supported earlier diagnosis (3). In the United States, AI is being deployed across urban and academic medical centers. Recent research from Standford University (4) has explored the end-to-end development of an automated system to deploy AI models based on clinician request in the EHR, and Stanford (4) and Vanderbilt (5), among others, are developing and evaluating systems that monitor deployed AI in real-time to identify concerns such as performance and fairness drift.

The current focus on AI deployments in urban areas and the history of US urban-rural digital divides raises concerns that the promise of AI may not be realized in rural communities. Slow or lack of AI adoption in rural communities may exacerbation well-documented health disparities, including higher mortality rates, diagnoses at more advanced stages of disease, delayed critical interventions, and longer inpatient stays. Without the benefits of AI-driven improvements in patient outcomes and increased efficiency, rural healthcare facilities may fall farther behind their urban counterparts and rural hospital closure rates may continue to rise. Deploying AI in rural healthcare facilities may also pose unique challenges to those being explored and addressed by the urban, academic medical centers serving as early AI adopters. Simply applying existing AI models from urban or academic medical centers alone may not be enough to provide rural areas with sustainable, equitable access to healthcare AI. First, AI models are known to not perform as well at when transferred to new sites with distinct patient populations (6). Moreover, AI models can change in performance over time (7,8). Given the relative sparsity of patients at any one rural site and the likelihood that these rural sites may not have the appropriate staff or resource, rural medical centers may not have capacity to localize models developed elsewhere, to train their own models, design locally responsive implementation strategies, or maintain models over time (9).

With 56 million people (∼18% of the US population) (10) residing in areas designated as rural or borderline rural by the Centers for Medicare and Medicaid Services (CMS)(11), a more thorough understanding of the current state and barriers to use of AI in rural care facilities in essential for the medical and public health communities to advance the health of rural populations and reduce geographic health disparities. We conducted a scoping literature review to answer the following research questions: For what tasks and with which techniques have AI been developed or evaluated in the rural US? What gaps, if any, exist with the application of AI in rural US healthcare? What challenges, if any, limit the development, implementation, or evaluation of AI in the rural US?

## Materials and Methods

### Search strategy and selection criteria

We searched PubMed, Embase, and WebOfScience for literature describing AI development or use at medical centers in the rural United States. Table 1 indicates the queries used to retrieve literature for review. We included peer-reviewed, original research studies indexed after January 1, 2010 and through our search date of April 29, 2025. Studies were required to discuss the development, implementation, or evaluation of AI tools in rural US healthcare, including frameworks that help facilitate AI development (e.g., data warehouses). Thus, we consider AI technologies that are developed in the rural US or developed outside the rural US and applied or validated in the rural US. We included both EHR-based implementations and non-EHR-based implementations.

**Table 1.** Search terms used to query PubMed, Embase, and WebOfScience and number of results retrieved from each database.

| Database | Search Term | # Results (before removing duplicates) |
| --- | --- | --- |
| PubMed | ("rural health"[MeSH Terms] OR "Rural Health Services"[MeSH Terms] OR "community medicine"[MeSH Terms] OR "rural population"[MeSH Terms] OR "community health centers"[MeSH Terms]) AND ("artificial intelligence"[MeSH Terms] OR "data science"[Text Word] OR "clinical decision support"[Text Word] OR "informatics"[MeSH Terms]) AND ("humans"[MeSH Terms]) AND (2010:2024[pdat]) | 1441 |
| Embase | ('rural health'/exp OR 'rural health care'/exp OR 'community medicine'/exp OR 'health center'/exp OR 'rural population'/exp) AND ('artificial intelligence'/exp OR 'data science'/exp OR 'clinical decision support system'/exp OR 'information science'/exp) AND 'human'/exp | 556 |
| WebOfScience | (TS="rural health" OR TS="Rural Health Services" OR TS="community medicine" OR TS='rural population' OR TS='community health centers') AND (WC="Computer Science, Artificial Intelligence" OR TS="data science" OR TS="clinical decision support" OR TS="informatics") | 766 |

After a pre-screening review of selected titles, we discovered two works – an editorial by Cecchetti (12) and research describing the development of longitudinal data including rural US (13) – that would likely be cited by titles relevant to this survey. Thus, any studies citing either were also included for screening.

We used Covidence (14) to facilitate the organization, title and abstract screening, and full-text screening of references. After removing duplicates, each abstract was screened for eligibility by both authors and discrepancies discussed for consensus. For studies with eligible abstracts, the full text was screened for eligibility by both authors, with reason for exclusion noted and discrepancies again discussed for consensus.

### Data extraction and analysis

For those studies deemed eligible after full text screening, we extracted relevant publication, geographic, clinical, and AI model information. We collected publication year and type (e.g., conference proceedings, journal article). We documented the state (if available) and geographic region (derived from state as necessary) of the first author’s institution, last author’s institution, origin of data, and location at which the AI tool was developed, evaluated, or implemented. We did not collect this information for external models evaluated as a baseline or comparison at a rural medical facility. We also collected information regarding the medical specialty and clinical task for which the AI was developed or applied.

In each study, we determined if the AI developed or evaluated was predictive (i.e., statistical model, machine learning model) or generative (e.g., large language model, image generation model) along with the type of algorithm (e.g., random forest, gradient boosted tree, neural network, GPT-3.5). We collected details on the evaluation strategy for the AI model and whether the work was an implementation study. We also determined which stage(s) of the AI lifecycle defined by De Silva and Alahakoon (15) where reported in the study. We noted the type of data used in the AI (e.g., structured EHR data, clinical text) and determined if the data was from a single rural medical center, multiple medical centers (e.g., across multiple institutions affiliated with an academic medical center), or a nationwide cohort (e.g., All of Us (16)). Finally, we collected any information related to limitations or hindrance to AI in rural healthcare as disclosed in the “Discussion” or “Limitations” sections of each paper.

We used Google Forms to facilitate consistent extraction of relevant details and exported all data for analysis in Python.

### Role of the funding source

The funder of the study had no role in study design, data collection, data analysis, data interpretation, or writing of the report.

## Results

Our search strategy found 2,792 studies, with 2,601 passing initial abstract screening and 26 studies meeting inclusion criteria after full text screening (Figure 1). The majority of studies (n=22, 85%) were journal articles and the remaining 15% (n=4) were from conference proceedings. Table 2 provides an overview of the 14 papers that discussed predictive AI models and Table 3 describes the 12 papers that discussed data or research infrastructure. No studies discussed generative AI models trained, evaluated, or deployed in a rural setting. Figure 2 shows how the literature evolved between 2012 to 2023. Between 2010 and 2013, research focused on predictive AI over data or research infrastructure. By 2015, there was an equal number of predictive modeling and infrastructure papers. Work in data and research infrastructure dominated this landscape between 2015 and 2021. During this time, total work in predictive AI for rural US health was initially stagnant (2014–2017) before seeing slight growth (2018–2021). In 2022, both predictive ML and infrastructure literature increased substantially, and from 2022, predictive AI studies again outnumbered infrastructure contributions.

**Figure 1.**
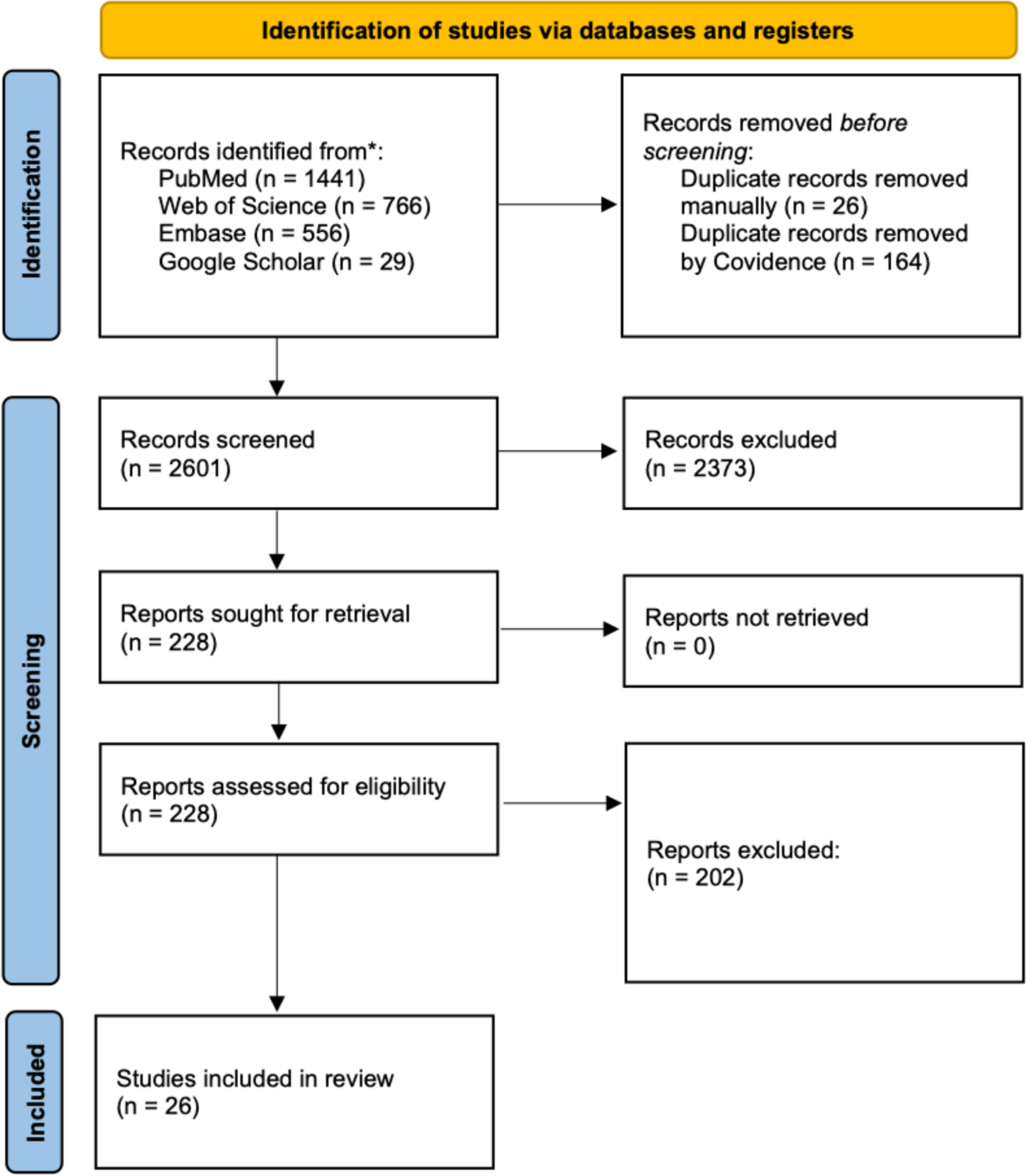
PRISMA scoping review flowchart illustrating the screening process.

**Figure 2.**
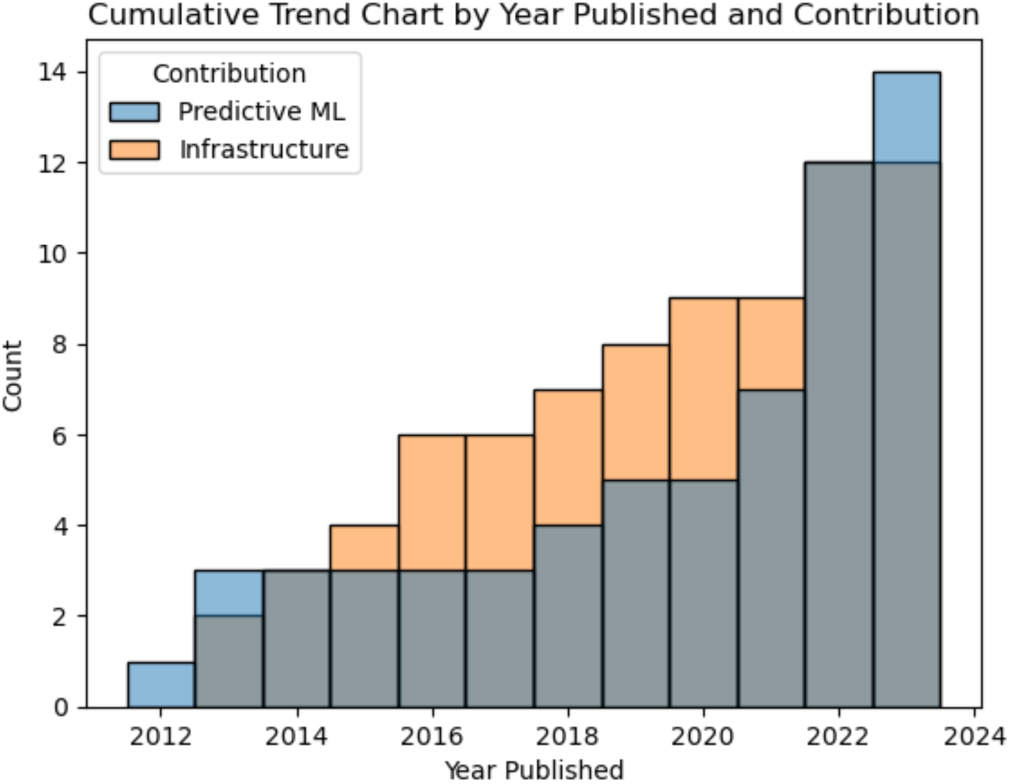
Cumulative trend chart comparing predictive machine learning and infrastructure contributions by year.

**Table 2.** Summary chart describing the data data collection for the predictive ML papers included in our review. Note: JA = Journal Article, CP = Conference Proceedings, PDB = publicly available, non-EHR database. Data denoted as other contains billing data, health department data or a combination of biological and environmental data.

|  | Year | Type | Data | ML Type | Validation Type | Metrics | Medical Specialty | Prediction Task | Lifecycle Stage |
| --- | --- | --- | --- | --- | --- | --- | --- | --- | --- |
| (34) | 2023 | JA | EHR | Tree Ensemble<br>Linear Regression | Holdout (1x) | C-statistic | Neurology | Predicting age of "young strokes" | Develop |
| (35) | 2022 | JA | EHR,<br>Clinical Text | Tree Ensemble<br>Gaussian Process<br>KNN<br>Unsupervised Learning | k-fold cross-validation | Accuracy<br>F1-Score<br>Recall<br>Precision<br>G-score<br>Balanced accuracy | COVID-19 | Predicting covid-19 test results based on testing reason | Design<br>Develop |
| (36) | 2022 | JA | EHR,<br>Clinical Text | Tree Ensemble<br>Non-tree ensemble<br>Logistic regression<br>Single DT | External Validation (Temporal), k-fold cross-validation | Accuracy<br>F1-Score<br>Recall<br>Precision<br>G-score<br>Balanced accuracy | Primary Care | Identify medical reason for patient appointment | Design<br>Develop |
| (17) | 2022 | JA | EHR | Logistic regression | Clinical trial | No quantitative ML metrics | Endocrinology | Pre-diabetes detection | Develop<br>Deploy |
| (37) | 2018 | CP | EHR,<br>Other | Logistic regression<br>Single DT | k-fold cross-validation | C-statistic | Obstetrics | Prediction of preterm birth | Design<br>Develop |
| (38) | 2022 | JA | PDB | Tree Ensemble<br>Single DT | None performed (Variable Importance) | No quantitative ML metrics | Oncology | Predicting breast cancer tumor stage at diagnosis by county | Design<br>Develop |
| (39) | 2013 | JA | EHR | Logistic regression | External Validation (Temporal) | TPR/Sensitivity /Recall<br>Specificity<br>PPV<br>NPV | Internal Medicine | Predict risk of 30 day readmission | Design<br>Develop |
| (18) | 2023 | JA | EHR | Logistic regression<br>Linear Regression | Clinical trial | Correlation between predicted/true events | Cardiology | Cardiovascular disease risk management | Deploy |
| (40) | 2021 | JA | EHR | Tree Ensemble | Holdout (1x) | Accuracy, F1-Score, Precision, TPR/Sensitivity /Recall, Specificity, C-statistic | COVID-19 | Predict extent of healthcare utilization by COVID positive patients | Design<br>Deploy |
| (41) | 2012 | JA | EHR | Logistic regression | External Validation (Geographic) |  | Trauma | massive blood transfusion protocol for trauma patients | Design<br>Develop |
| (42) | 2019 | JA | PDB | Unsupervised Learning | None performed | N/A | Operations | Calculating efficiency of the provision of a single health service | Design |
| (25) | 2021 | JA | Other | Deep Learning (LSTM) | External Validation (Temporal) | Discounted cumulative grain (DCG) | COVID-19 | Recommend counties of increased incidences for intervention | Design<br>Develop<br>Deploy |
| (43) | 2022 | JA | EHR | Tree Ensemble | Holdout (1x) | Root mean squared error<br>Pearson's correlation | Orthopedic Surgery/LOS Prediction | LOS prediction after total joint replacement including hip, knee, and shoulder | Design<br>Develop |
| (44) | 2013 | CP | EHR,<br>Other | Tree Ensemble | Holdout (bootstrapped) | C-statistic | Cardiology | 30-day readmission for heart failure patients | Design<br>Develop |

**Table 3.**
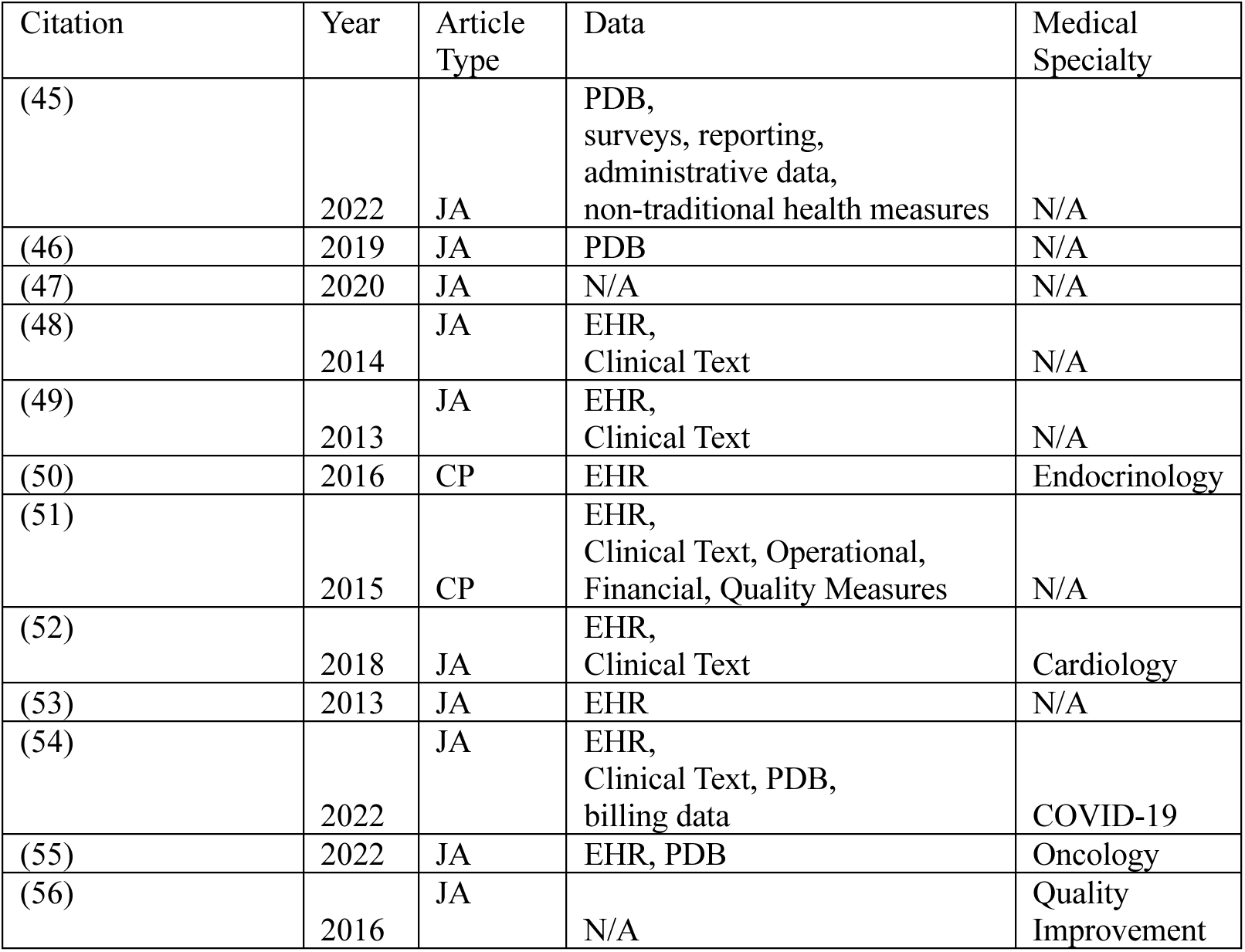
Summary chart describing the data data collection for the infrastructure papers included in our review. Note: JA = Journal Article, CP = Conference Proceedings, PDB = Public, non-EHR database,

We also examined the medical specialty for both predictive modeling and infrastructure papers. For studies of predictive AI, models were commonly related to the COVID-19 pandemic (n=3, ∼21% of predictive AI subset, 4% total) and cardiology outcomes (n=2, ∼14% of predictive AI subset, 8% total). The remaining 9 (64% of predictive AI subset, 35% total) explored a wide variety of clinical settings, including endocrinology, internal medicine, neurology, obstetrics, oncology, primary care, trauma, orthopedic surgery, and hospital operations. Half of the infrastructure papers (n=6, 23% total) were not concerned with a single specialty but focused on assimilating EHR data from inpatient and outpatient settings. The remaining infrastructure papers included tools focused on supporting analyses for COVID-19, cardiology, endocrinology, oncology, public health, and quality improvement.

Studies of AI in rural health included researchers and communities from across the US (see Figure 3), highlighting broad interested in AI across diverse rural communities. Approximately 8% (n=2) of studies did not list the state or US region in which data were collected and one study included data from Puerto Rico – a US territory. Of the papers that contributed the development or validation of a predictive AI models, approximately 79% (n=11, 42% total) were authors by researchers from the same state from which some or all of the data originated.

**Figure 3.**
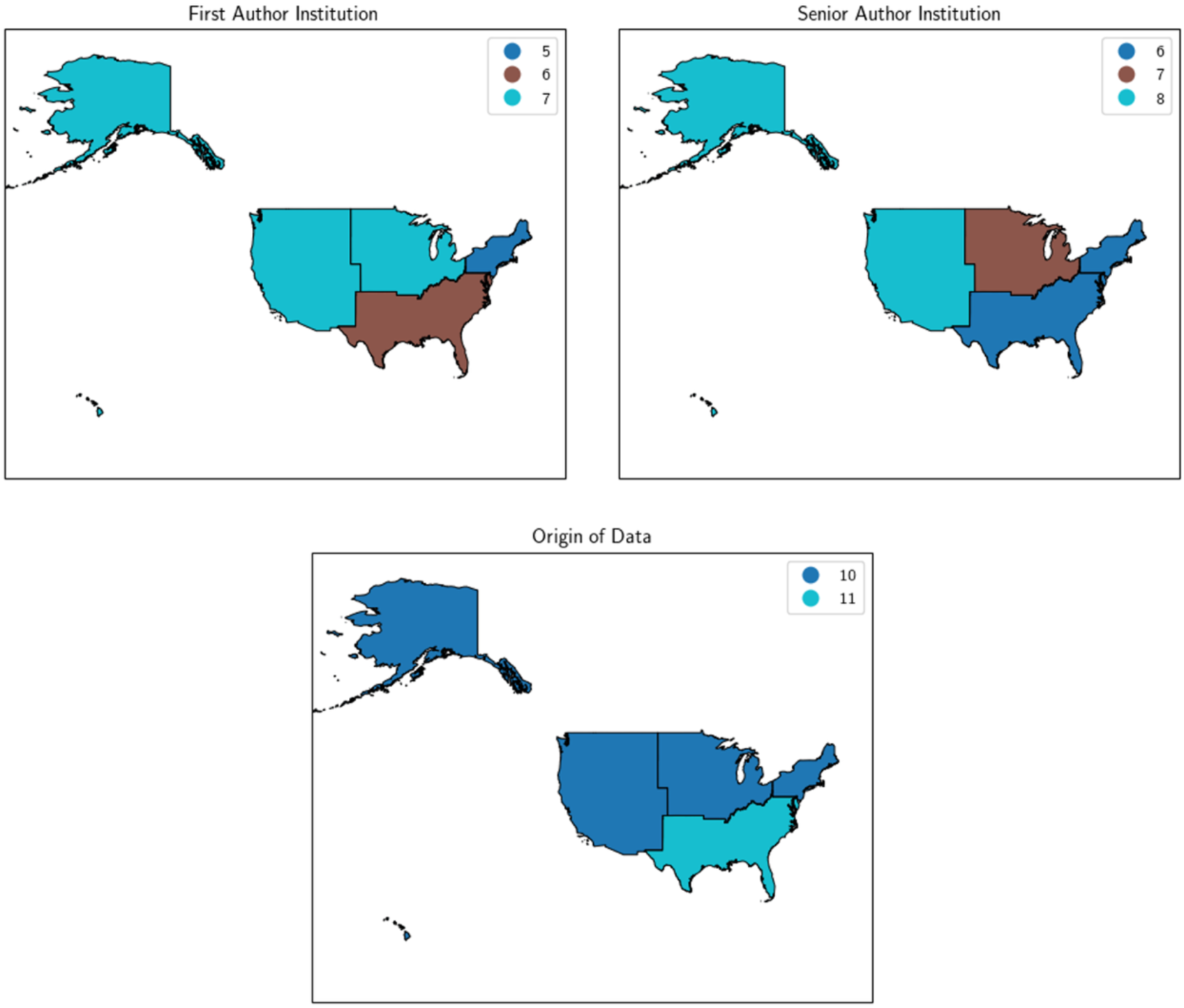
Geographic heatmaps of first author, senior author, and data origin per paper.

### Predictive AI models

The most common predictive algorithms were tree ensembles such as gradient-boosted trees or random forests (n=7, 50% of predictive AI subset). Logistic regression (n=6, 43%) and non-ensemble decision trees (n=3, 21%) were the next most frequent, followed by linear regression (n=2, 14%), and various forms of unsupervised learning (n=2, 14%). Adaboost (a non-tree-based ensemble), deep learning in the form of long short-term memory (LSTM) networks, Gaussian processes, and k-nearest neighbors were each used in a single study. Three-fourths of studies (79%, n=11) conducted model validation, one study pursued multiple forms of model validation, and two studies did not present validations. The most common validation procedures (n=3 papers each, 21%) were temporal data splitting, random data splitting with a single holdout test set, and k-fold cross-validation. Two papers (14%) evaluated a predictive model as part of a clinical trial. Finally, one paper (7%) performed geographic external validation by sampling validation points from a different geographic region location. The most common validation metric was binary area under the receiver operating characteristic curve (AUROC, equiv. C-statistic).

Of the papers discussing predictive models, almost 80% (n=11) used structured EHR data. The next most common data source was clinical narrative text (e.g., notes and clinical summaries) and publicly available, non-EHR data sources, each being used in 2 papers (14%). One paper each (7%) used billing data, health department data or a combination of biological and environmental data. In approximately 71% of predictive modeling papers (n=10), data originated from a resource combining information from multiple rural medical providers, and the remaining 29% (n=4) used data from a single rural medical centers. Approximately 29% of papers (n=4) used data originating from an independent, non-academic medical center. Approximately 14% (n=2) used data from an academic medical center with rural satellite sites or data not associated with a medical center (i.e., external data sources/publicly available datasets). The remaining 57% (n=8) of predictive AI papers used data not associated with a single medical center.

We consolidated the 19 detailed stages of the proposed AI lifecycle (15) into three broad categories: design (i.e., related to problem formulation and data acquisition), develop (i.e., related to model develop and initial evaluation), and deploy (i.e., model deployment, evaluation, and monitoring). Of the predictive AI papers, 79% (n=11) contained an aspect of the design phase, 79% (n=11) contained an aspect of the develop phase, and 29% (n=4) contained an aspect of the deploy phase. Two papers (14%) could be considered as implementation studies. Both implementation studies (17,18) noted difficulties with adoption of the AI tools due to reduced financial and technical resources.

### Infrastructure

Our review highlighted several initiatives that could support AI development and dissemination in rural healthcare settings, such as the Appalachian Informatics Platform. Most such studies were associated with the western US (n=7, ∼63%). The majority of infrastructure papers (11 of 12, 92%) accessed structured EHR as the primary data source. Two papers included publicly available data or clinical narrative text (18% each), and one paper (9%) accessed data from a health information exchange, including public health departments laboratory data, billing, operational, and quality measures in their database. All papers used data combined across multiple sources.

All infrastructure papers focused on data collection. Data harmonization (n=9, 75%) was a common concern and half of the infrastructure papers (n=6) sought to provide data visualization tools. One paper (8%) aimed to enable data sharing within the infrastructure. Most papers (n=9, 75%) did not explicitly mention support for AI as a current or future use case of the infrastructure. Two infrastructure papers (n = 2) included tools to support predictive analytics and one paper demonstrated the development of an AI model. Two papers described tools that explicitly noted AI modeling as a future direction for the infrastructure tools.

### Challenges

Table 4 presents challenges for AI development and deployment in rural settings as identified in reviewed studies. The most common acknowledged challenge was a lack of reliable, high quality data sources and small data volumes. A lack of data science expertise was also commonly highlighted in the studies. The existing urban-rural healthcare divide, lack of data harmonization and low data quality, and lack of technical infrastructure were also noted. Other challenges included differences in disease prevalence and patient demographics across geographic regions, difficulties in organizational/community engagement, lack of medical staff or medical staff training in informatics tools, and the need for AI governance.

**Table 4.** Distribution of challenges present in each paper covered in our scoping review.

| Challenge | Number of papers (%) |
| --- | --- |
| Lack of reliable data sources/volume | 13 (50%) |
| Lack of data science expertise | 8 (30.77%) |
| Existing urban-rural healthcare divide | 7 (26.92%) |
| Lack of data harmonization | 7 (26.92%) |
| Lack of technical infrastructure | 7 (26.92%) |
| Differences in disease prevalence | 5 (19.23%) |
| Differences in demographics | 4 (15.38%) |
| Difficulties in organizational/community engagement | 4 (15.38%) |
| Lack of work evaluating clinical prediction models to rural communities | 3 (11.54%) |
| Lack of staff or staff training | 2 (7.69%) |
| Need for AI governance | 2 (7.69%) |
| Lack of non-data science, non-clinical expertise | 1 (3.85%) |

## Discussion

Research on AI solutions for rural healthcare mirrors the adoption of EHR systems in the rural US. After an initial surge in research in 2013 and 2014, new contributions in AI development slowed until 2022 and then increased, in part, as a response to the coronavirus pandemic. This turning point in AI/ML development can be compared to how the HITECH Act (19) required the adoption and use of EHR systems in American healthcare systems – including in the rural US – and as a result is hypothesized to be a contributing factor in EHR adoption in the rural US (20). Additionally, rural hospitals may have invested limited technology resources into telehealth solutions (21) which may be contributing to a delay in AI investment.

For predictive AI models, applications most commonly targeted resource allocation and distribution. We noted several attempts to predict resource utilization surrounding COVID-19 testing needs and case distributions. This makes sense as smaller medical centers could quickly be overwhelmed by surging case loads, especially given limited staffing, making models to predict where public health agencies could efficiently direct resources in rural communities imperative. However, we noted few AI solutions for acute medical events faced by rural patients, such as trauma and stroke. Outcomes are worse for rural patients suffering from an acute neurological event or trauma (22,23) and as such these conditions pose an opportunity for AI to improve care for rural patients. The limited availability of time-critical specialties such as trauma/emergency medicine, neurology, and cardiology in rural areas often necessitates patients with such conditions be transferred to larger, more resourced hospitals. We posit that it is possible to improve health outcomes for these patients by developing and evaluating targeted AI models for these scenarios. Our review indicates a gap and highlights opportunity for innovation in leveraging AI tools to predict and support patient transfers in rural communities.

Practical limitations may be influencing and limiting the types of AI models evaluated in rural US medical facilities. The most frequent model employed, tree-based ensembles such as random forests and gradient-boosting trees, are common and powerful algorithms that do not require the same levels of energy or computational overhead as neural networks and large language models (24). This means such models may be more feasible for implementation at rural medical centers where computational resources may be limited. We observed a limited exploration of deep learning and advanced neural network models, including generative AI such as large language models, in rural health care settings. Deep learning was utilized in one paper to predict counties in West Virginia likely to face an increase in COVID-19 infections using non-text epidemiological data (25). No generative AI models were captured in our review. We also did not find studies translating advances in AI-based pathology and radiology diagnostic tools to rural communities. These well-researched and successful use cases of deep learning (3,26–28) could be particularly useful at rural medical centers with limited specialty providers and support more efficient remote care. One reason for this lack of deep learning use cases in our review may be that such specialized deep learning models require intensive and expensive computational power, which may be infeasible for small, rural medical centers, many of which are financially tenuous and lack the ability to invest in computational resources. We note this lack of research into deep learning for rural US healthcare has introduced a rural-urban divide in AI technologies – widening the existing rural-urban healthcare divide. Unfortunately, this divide is likely to expand if research into generative AI does not include evaluating performance for rural US healthcare and improving accessibility to underserved communities.

Our review highlighted few studies of AI moving beyond the design and develop stages, leaving a clear gap in our understanding of how to deploy and sustain AI models in rural settings. Several challenges noted in the reviewed studies may provide insight into this lack of translation from research to implementation. Multiple papers noted a lack of technical infrastructure and dedicated staff to train and validate AI solutions. This may be attributed to a lack of funding, which was another common barrier to adoption. Limited data resources and sample sizes for training and evaluating complex models and measuring the impact of deployed tools also poses a unique challenge in rural health care facilities. Further research is needed to enhance the translation of state-of-the-art modeling techniques into effective AI tools for use in the rural US, including exploring partnerships between academic medical centers and rural communities and solutions to logistic challenges of such partnerships, including data and resource sharing.

Validation of AI tools in the rural US was underwhelming, and ultimately, neglected. The most common form of model validation employed was a single random holdout test set. In this paradigm, a majority of available data is used for model training while the remaining, unseen data is reserved for evaluation. This approach can provide overly optimistic indications of model performance and obscure indications of model overfitting if the test set by random chance provides an advantageous or disadvantageous split between training and validation data (29). This may be of particular concern for small datasets from rural facilities where test sets are small and subject to high variability. Only one paper used multiple holdouts via bootstrapping, and three papers use a form of k-fold cross-validation. These techniques train and evaluate models multiple times using random partitions of the data and help provide more realistic assessments of model performance; however, these approaches may not address geographic or temporal generalizability. External validation is always recommended and was considered in four papers that performed multi-site external evaluation or temporal splitting for validation.

Reviewed studies highlighted a lack of reliable data sources or limited data volume as a potential challenge in developing and adopting AI. Patient-level EHR data was often limited to specific medical centers, which can only provide small sample sizes in rural communities. While existing patient-level EHR databases such as All of Us (16) or electronic ICU (eICU) (30) contain proxies for rurality such as most frequent ZIP-3 codes per site or site size, these sources are not widely used for research in AI for the rural US. Moreover, these databases may not reflect demographic or medical event prevalence of a specific rural area, a widely noted concern with model development and evaluation (31). Synthetic data generation (32) and federated learning (33) are two technical approaches that could help mitigate these sample size and data representativeness concerns, but such approaches have yet to be applied to support AI in rural health and may require additional computational and analytic staff support.

## Conclusion

Rural medical centers are overburdened and understaffed, making the promise of efficiency and improved care quality through AI tools particularly critical. In this work, we performed a scoping review of healthcare AI tools and infrastructure in the rural United States. Most research focused on predictive modelling with EHR data, commonly using ensembles of tree-based models, decision trees, and regression techniques. The rural US faces challenges in data volume and quality, leading to less robust evaluation of predictive models. The lack of technical infrastructure, data science staffing, and funding have led to a growing urban-rural divide in AI research. Narrowing this divide is a growing necessity, especially with the rise of generative AI and the risk of further exacerbating the urban-rural divide if effective generative AI tools are broadly available to all communities. Given the limited research into rural use of clinical AI and the challenges to deploying AI in rural settings, research and operations institutional partnerships, as well as policy initiatives, will be necessary to realize the promise of healthcare AI for all individuals and communities across the United States.

## Data Availability

All data produced in the present study are available upon reasonable request to the authors

## Contributors

KEB conceived of the study, managed search and data extraction, reviewed all studies, extracted data from included studies, and drafted the initial manuscript. SED reviewed all studies, extracted data from included studies, revised the manuscript, and provided critical appraisal before submission of its final version. All authors had full access to all the data in the study and had final responsibility for the decision to submit for publication.

## Conflicts of Interest

All authors declare no financial or non-financial competing interests.

## Acknowledgements

This study was supported by the National Institutes of Health (grant number T15LM007450). The funding agency was not involved in the design, conduct, or reporting of this study.

## Notes

### Competing Interest Statement

The authors have declared no competing interest.

