## Supplemental Materials for "Gaps in Artificial Intelligence Research for Rural Health in the United States: A Scoping Review"

Keywords:

### S1. Analysis of ACS Data Regarding Rural Health

While machine learning in healthcare has made massive strides in recent years, most progress is typically the result of EHR data from large, primarily urban academic medical centers. There is, however, a significant proportion of the US population that resides in a rural area. Using the American Community Survey (ACS, 2018-2022)(1), we calculated that approximately 56,000,000 people (~18% of the US population) reside in a ZIP Code Tabulation Area (ZCTA) designated as rural or borderline rural by the Centers for Medicare and Medicaid Services (CMS)(2). Rural or borderline rural areas comprise about 80% of US land area when calculating cumulative land size of ZCTAs in the CMS ambulance fee schedule. We refer to areas with a rural or borderline rural distinction in the fee schedule as 'not urban' for proceeding analysis. All analysis is performed using the R statistical computing software, and all statistical tests are Welch's two sample t-test.

We can further use the ACS to perform further comparisons. First, we consider Area Disadvantage Index (ADI) for the ZCTAs in the CMS ambulance fee schedule. We use Berg et al.'s reformulation of ADI (ADI-3) which is composed of components measuring Financial Strength, Economic Hardship and Inequality, and Educational Attainment(3). Some ZCTAs may not have had these values defined. Thus, missing values are dropped from our analysis.

Table S1. Area deprivation index (ADI) and individual components for urban and not urban populations. Not urban is equivalent to rural and borderline rural areas. Bold indicates least disadvantaged value. Italics indicates most disadvantaged values.

| Population | ADI (higher indicates more disadvantage) | Financial Strength (higher is better) | Economic Hardship/Inequality (lower is better) | Educational Attainment (higher is better) |
|---|---|---|---|---|
| Urban | **95.17 (94.83-95.52)** | **106.97 (106.61-107.33)** | **99.3 (98.97-99.63)** | **100.43 (100.1-100.76)** |
| Not Urban | 105.87 (105.62-106.13) | 91.95 (91.74-92.15) | 101.22 (100.92-101.52) | 98.8 (98.48-99.12) |
| Rural | *106.63 (106.32-106.95)* | *91.24 (90.99-91.49)* | *101.47 (101.09-101.84)* | *97.97 (97.56-98.38)* |
| Borderline Rural | 104.85 (104.43-105.26) | 92.9 (92.56-93.25) | 100.9 (100.4-101.39) | 99.92 (99.42-100.41) |

Table S1 gives ADI values and values of relevant factors of ADI including Financial Strength, Economic Hardship/Inequality, and Educational Attainment. Consistently across variables, urban areas had the most advantage, and rural areas had the least advantage.  Using Welch's T-Test, the difference in means between not urban and urban populations is statistically significance across all variables (p < 2.2e-16 for ADI, FS, EHI and p=1.47e-12 for EA). This indicates that there is a small, yet significant difference between not urban and urban populations with not urban areas having a slightly more financial disadvantage compared to urban populations.

Next, we consider poverty level for ZCTAs partitioned by the CMS ambulance fee schedule using the ACS data regarding poverty status in the past 12 months (Table S1701). As before, missing values are dropped from our analysis. Urban areas had an average of 12.35% (CI 12.17-12.54%) of residents living below the poverty level. Not urban areas had an average of 14.59% (CI 14.4-14.78%) of residents below the poverty level. Rural and borderline rural areas had averages of 14.66% (14.42-14.9%) and 14.5% (14.2-14.81%) of residents below the poverty level, respectively. We performed Welch's T-Test (assuming normality since our smallest sample size was >7,000) and found that the 2.24% (p < 2.2e-16) difference between Not Urban and Urban areas is statistically significant. Welch's T-Test found that the <1% difference between rural and borderline rural was not statistically significant.

Table S2. Percentage of urban and not urban population with and without insurance.

| Population | With Insurance | | Without Insurance |
|---|---|---|---|
| | 1 Insurance | 2+ Insurances | |
| Urban | 74.02 (73.85-74.19) | 17.99 (17.84-18.14) | 7.99 (7.87-8.11) |
| Not Urban | 68.76 (68.56-68.96) | 21.9 (21.72-22.08) | 9.34 (9.2-9.48) |
| Rural | 69.83 (69.58-70.08) | 21.56 (21.34-21.78) | 8.61 (8.45-8.77) |
| Borderline Rural | 67.32 (66.99-67.65) | 22.36 (22.06-22.66) | 10.32 (10.08-10.56) |

Next, we analyzed insurance status (ACS table B27010) for the populations defined by the CMS ambulance fee schedule. First, in Table S2, we consider proportion of each population with one type of insurance coverage, two or more types of insurance coverage, or no insurance. The urban population (7.99% CI: 7.97-8.00%) has the smallest proportion of uninsured with borderline rural populations (10.32% CI: 10.29-10.35%) having the highest rates of being uninsured. Comparing urban and not urban populations, we see a statistically significant 1.35% (p < 2.2e-16) difference in uninsurance rates.

We can further analyze the insured population by considering the proportion of the population with only one type of insurance and the proportion of the population with two or more different types of coverage. From the ACS data, we define private insurance as direct purchase or employer-based and government-sponsored as Medicare, Medicaid, and Tricare or VA Health Care.

Table {1ins-comp}. Percentage of urban and not urban population with one type of insurance coverage with private or government sponsored insurance.

| Population | Private Health Insurance (%) | Gov. Sponsored Health Insurance (%) |
|---|---|---|
| Urban | 70.27 (70.24-70.31) | 29.73 (29.69-29.76) |
| Not Urban | 62.98 (62.94-63.02) | 37.02 (36.98-37.06) |
| Rural | 63.24 (63.2-63.29) | 36.76 (36.71-36.8) |
| Borderline Rural | 62.62 (62.56-62.68) | 37.38 (37.32-37.44) |

From Table S2, we see that the urban population (74.02% CI: 73.85-74.19%) is the population with the highest insured with one type of insurance. The difference between the percentages of urban and not urban populations with only one type of insurance is statistically significant at 5.26% ($p < 2.2e-16$). We can examine how the population with one insurance type is distributed across private insurance and government-sponsored insurance in Table {1ins-comp}. Of those with only one insurance plan, the urban population (70.27% CI: 70.24-70.31%) had the highest percentage of those covered with private health insurance (and the lowest with government-sponsored health insurance), and the borderline rural population had the highest percentage (37.38% CI: 37.32-37.44%) of coverage by government-sponsored health insurance (and the lowest with private health insurance). The difference between the percentages of urban and not urban population is 7.29% ($p < 2.2e-16$).

Table S3. Percentage of urban and not urban population with more than one insurance coverage with private or government sponsored insurance

| Population | Private Only | Gov. Sponsored Only | Combination of Private and Gov. Sponsored |
|---|---|---|---|
| Urban | 11.99 (11.97-12.02) | 15.78 (15.75-15.81) | 72.23 (72.2-72.26) |
| Not Urban | 7.43 (7.41-7.45) | 18.09 (18.06-18.12) | 74.48 (74.44-74.51) |
| Rural | 7.5 (7.47-7.52) | 18.09 (18.05-18.13) | 74.41 (74.36-74.45) |
| Borderline Rural | 7.34 (7.31-7.38) | 18.09 (18.03-18.14) | 74.57 (0.7451-0.7463) |

Interestingly, the borderline rural population (22.36% CI: 22.06-22.66%) have the highest percentage with at least two types of insurance coverage, and the urban population has the lowest percentage (17.99% CI: 17.84-18.14%). Between urban and not urban populations, the difference between the percentage of not urban and urban populations with at least two different types of insurance is 3.91% ($p < 2.2e-16$). Table S3 provides the distribution of possible combinations being private only, government-sponsored only, or a mix of private and government sponsored. We see that the urban population (11.99% CI: 11.97-12.02%) has the highest percentage of those with at least two insurance types with both types being private insurance. Similarly, the not urban and borderline rural populations have the highest percentage of having both types of insurance plans being government-sponsored insurance (18.09 CI: 18.06-18.12%) or a combination of private and government-sponsored (74.57% CI: 74.57-74.63%), respectively.

**S2. Preferred Reporting Items for Systematic reviews and Meta-Analyses extension for Scoping Reviews (PRISMA-ScR) Checklist**

Table S4. PRISMA-ScR checklist for our literature review.

| SECTION | ITEM | PRISMA-ScR CHECKLIST ITEM | REPORTED ON PAGE # |
|---|---|---|---|
| **TITLE** | | | |
| Title | 1 | Identify the report as a scoping review. | 1 |
| **ABSTRACT** | | | |
| Structured summary | 2 | Provide a structured summary that includes (as applicable): background, objectives, eligibility criteria, sources of evidence, charting methods, results, and conclusions that relate to the review questions and objectives. | 2 |
| **INTRODUCTION** | | | |
| Rationale | 3 | Describe the rationale for the review in the context of what is already known. Explain why the review questions/objectives lend themselves to a scoping review approach. | 2-3 |
| Objectives | 4 | Provide an explicit statement of the questions and objectives being addressed with reference to their key elements (e.g., population or participants, concepts, and context) or other relevant key elements used to conceptualize the review questions and/or objectives. | 3 |
| **METHODS** | | | |
| Protocol and registration | 5 | Indicate whether a review protocol exists; state if and where it can be accessed (e.g., a Web address); and if available, provide registration information, including the registration number. | 6 |
| Eligibility criteria | 6 | Specify characteristics of the sources of evidence used as eligibility criteria (e.g., years considered, language, and publication status), and provide a rationale. | 5 |
| Information sources* | 7 | Describe all information sources in the search (e.g., databases with dates of coverage and contact with authors to identify additional sources), as well as the date the most recent search was executed. | 5 |
| Search | 8 | Present the full electronic search strategy for at least 1 database, including any limits used, such that it could be repeated. | 5, 19 |
| Selection of sources of evidence† | 9 | State the process for selecting sources of evidence (i.e., screening and eligibility) included in the scoping review. | 5-6 |
| Data charting process‡ | 10 | Describe the methods of charting data from the included sources of evidence (e.g., calibrated forms or forms that have been tested by the team before their use, and whether data charting was done independently or in duplicate) and any processes for obtaining and confirming data from investigators. | 6 |
| Data items | 11 | List and define all variables for which data were sought and any assumptions and simplifications made. | 6 |
| Critical appraisal of individual sources of evidence§ | 12 | If done, provide a rationale for conducting a critical appraisal of included sources of evidence; describe | Click here to enter text. |

| SECTION | ITEM | PRISMA-ScR CHECKLIST ITEM | REPORTED ON PAGE # |
|---|---|---|---|
| | | the methods used and how this information was used in any data synthesis (if appropriate). | |
| Synthesis of results | 13 | Describe the methods of handling and summarizing the data that were charted. | 6 |
| **RESULTS** | | | |
| Selection of sources of evidence | 14 | Give numbers of sources of evidence screened, assessed for eligibility, and included in the review, with reasons for exclusions at each stage, ideally using a flow diagram. | 7 |
| Characteristics of sources of evidence | 15 | For each source of evidence, present characteristics for which data were charted and provide the citations. | 7-9, 22-23 |
| Critical appraisal within sources of evidence | 16 | If done, present data on critical appraisal of included sources of evidence (see item 12). | Click here to enter text. |
| Results of individual sources of evidence | 17 | For each included source of evidence, present the relevant data that were charted that relate to the review questions and objectives. | 7-9, 22-23 |
| Synthesis of results | 18 | Summarize and/or present the charting results as they relate to the review questions and objectives. | 7-9 |
| **DISCUSSION** | | | |
| Summary of evidence | 19 | Summarize the main results (including an overview of concepts, themes, and types of evidence available), link to the review questions and objectives, and consider the relevance to key groups. | 10-12 |
| Limitations | 20 | Discuss the limitations of the scoping review process. | 13 |
| Conclusions | 21 | Provide a general interpretation of the results with respect to the review questions and objectives, as well as potential implications and/or next steps. | 13 |
| **FUNDING** | | | |
| Funding | 22 | Describe sources of funding for the included sources of evidence, as well as sources of funding for the scoping review. Describe the role of the funders of the scoping review. | 7, 13 |
